# Febrile Seizure Risk Following Monovalent COVID-19 mRNA Vaccination in US Children Aged 2-5 Years

**DOI:** 10.1101/2024.03.12.24304127

**Authors:** Steven A Anderson, Elizabeth R Smith, Zhiruo Wan, Kandace L Amend, Alex Secora, Djeneba Audrey Djibo, Kamran Kazemi, Jennifer Song, Lauren E Parlett, John D Seeger, Nandini Selvam, Cheryl N McMahill-Walraven, Mao Hu, Yoganand Chillarige, Richard A Forshee

## Abstract

**Objective:** To evaluate febrile seizure risk following monovalent COVID-19 mRNA vaccination among children aged 2-5 years.

**Methods:** The primary analysis evaluated children who had a febrile seizure outcome in the 0-1 days following COVID-19 vaccination. A self-controlled case series analysis was performed in three commercial insurance databases to compare the risk of seizure in the risk interval (0-1 days) to a control interval (8-63 days).

The exposure of interest was receipt of dose 1 and/or dose 2 of monovalent COVID-19 mRNA vaccinations.

The primary outcome was febrile seizure (0-1 day risk interval).

A conditional Poisson regression model was used to compare outcome rates in risk and control intervals and estimate incidence rate ratios (IRR) and 95% confidence intervals (CIs). Meta-analyses were used to pool results across databases.

**Results:** The primary meta-analysis found a statistically significant increased incidence of febrile seizure, in the 0-1 days following mRNA-1273 vaccination compared to the control interval (IRR: 2.52, 95% CI: 1.35 to 4.69, risk difference (RD)/100,000 doses = 3.22 (95%CI -0.31 to 6.75)). For the BNT162b2 vaccination, the IRR was elevated but not statistically significant (IRR: 1.41, 95%CI: 0.48 to 4.11, RD/100,000 doses = -0.25 (95%CI -2.75 to 2.24).

**Conclusions:** Among children aged 2-5 years, the analysis showed a small elevated incidence rate ratio of febrile seizures in the 0-1 days following the mRNA-1273 vaccination. Based on the current body of scientific evidence, the safety profile of the monovalent mRNA vaccines remains favorable for use in young children.

**Article Summary:** In this self-controlled case series study, researchers evaluated whether there is an elevated risk of febrile seizure immediately following monovalent COVID-19 mRNA vaccination.

**What’s Known on This Subject:** The United States Food and Drug Administration previously noted a potential safety concern for seizure in children aged 2-5 years receiving the ancestral monovalent COVID-19 mRNA vaccines.

**What This Study Adds:** In this self-controlled case series that included participants aged 2-5 years from three commercial insurance databases, the incidence rate ratio of febrile seizures was significantly elevated in the 0-1 days following mRNA-1273 administration. Absolute risk was small.

## INTRODUCTION

On June 17, 2022 the United States (U.S.) Food and Drug Administration (FDA) granted emergency use authorization (EUA) for the monovalent COVID-19 mRNA vaccines developed by Pfizer-BioNTech (BNT162b2) for use in individuals aged 6 months through 4 years, and Moderna (mRNA-1273) for use in individuals aged 6 months through 17 years.^1^ The BNT162b2 vaccine was authorized as a three-dose primary series, and the mRNA-1273 vaccine was authorized as a two-dose primary series in young children. As of April 18, 2023 these initial formulations of the mRNA vaccines are no longer available for use and have been replaced by updated formulations based on currently circulating variants.^2^

Fever was one of the commonly observed systemic adverse events (SAEs) in children in the clinical trials of both mRNA vaccines, and in some instances, may result in febrile seizures. In clinical trials of mRNA-1273 vaccinations among individuals aged 24-36 months, ^3^ fever (≥38.0°C) was observed in 11.3% and 18.9% of participants following dose 1 and 2, respectively. Grade 3 fever (39.6°C-40.0°C) was observed in 0.3% and 1.2% of participants following dose 1 and 2, respectively, and grade 4 fever (>40.0°C) was observed in 0.3% and 0.3% of participants following dose 1 and 2. Fever was the most common grade 3/4 SAE observed in individuals aged 37 months – 5 years. Fever (≥38.0°C) was observed in 7.7% and 16.0% of participants aged 37 months – 5 years following dose 1 and 2, respectively. Grade 3 fever (39.0°C-40.0°C) was observed in 1.1% and 2.9% of participants following dose 1 and 2, and grade 4 fever (>40.0°C) was observed in 0% and 0.2% of participants following dose 1 and 2.

For BNT162b2, the most common severe systemic reaction among individuals aged 2-4 years was fever.^4^ Fever (≥38.0°C) was observed in 5.2%, 4.9% and 5.1% of participants following dose 1, 2, and 3 respectively. Fever (>38.9°C to 40.0°C) was observed in 0.7%, 1.1%, and 0.7% of participants following dose 1, 2, and 3 respectively, and fever (>40.0°C) was observed in 0.1%, 0.1%, and 0% of participants following dose 1, 2, and 3.

In a prior safety monitoring study, the FDA Center for Biologics Evaluation and Research (CBER) conducted near real-time surveillance (NRS) of the authorized COVID-19 vaccines in the pediatric population.^5,6^ NRS allows for the rapid identification of statistical signals for potentially elevated risk of evaluated outcomes following vaccination. Of the 19 outcomes evaluated for increased risk following exposure to the monovalent COVID-19 vaccines, the NRS detected a statistical signal for seizures/convulsions across the different doses (dose 1 and/or dose 2) following exposure to BNT162b2 in individuals aged 2-4 years and mRNA-1273 in individuals aged 2-5 years. Further evaluation of the potential signal was necessary as NRS was designed to be sensitive but not specific for screening and detection purposes.

The NRS used a broad outcome definition for seizures/convulsions and evaluated a 0-7 day risk interval. However, further examination of the data used in the NRS study indicated that most of the identified seizures/convulsions cases were febrile seizures. Accordingly, this current study uses the febrile case definition for children under the age of 5 years and a risk interval of 0-1 days, as used in prior FDA studies, to ensure that febrile cases are more likely to be associated with vaccination rather than other causes.^7^ This manuscript summarizes results of a self-controlled case series (SCCS) study conducted in 3 commercial insurance databases in the pediatric population aged 2-5 years. The primary objective was to determine the risk of febrile seizure in the 0-1 days following vaccination.

## METHODS

### Data Sources

This study used commercial health claims data from the Carelon Research, CVS Health, and Optum databases to capture COVID-19 vaccination data and medical claims data relevant to the outcomes of interest. COVID-19 vaccination data from participating Immunization Information System (IIS) jurisdictions were used to supplement claims data to improve capture of COVID-19 vaccinations (eTable 1). ^8^

### Study Population, Study Period, and Study Design

The study population consisted of enrollees aged 2-4 years who received a BNT162b2 COVID-19 vaccination and enrollees aged 2-5 years who received a mRNA-1273 COVID-19 vaccination. To be included in the study population, individuals were required to be continuously enrolled in the health plan at least 42 days prior to receiving the vaccine dose of interest.

Individuals were excluded if they had COVID-19 vaccination patterns that did not conform to expectations for the analysis of interest, such as subsequent doses occurring fewer than 3 days apart or receipt of more doses than approved. The start of the study period was June 17, 2022, the EUA for COVID-19 vaccinations in these age groups. The study end date was database specific (eTable 1).

We used a SCCS study design to compare the incidence of seizure outcomes following monovalent COVID-19 pediatric vaccine administration within pre-specified risk intervals (primary outcome: febrile seizures, 0-1 days; secondary outcomes: febrile seizures, 0-7 days and seizures/convulsions, 0-7 days) to a control interval (day 8 through the end of the 63 day observation period). In the primary outcome analysis, we included a washout period (days 2-7) prior to the control interval; for secondary outcomes, this period was included in the risk interval.

### Exposures and Outcomes

The primary exposure of interest was the receipt of dose 1 and/or dose 2 of the monovalent COVID-19 BNT162b2 (individuals aged 2-4 years) or mRNA-1273 (individuals aged 2-5 years) pediatric vaccines. Vaccinations were identified in administrative claims data through vaccine-specific codes including Current Procedural Terminology (CPT)/Healthcare Common Procedure Coding System (HCPCS) codes and National Drug Codes (NDCs) in the professional, outpatient, inpatient, or pharmacy care settings, as well as CVX (vaccine administered codes) in IIS data (eTable 2).

The primary outcome of interest was febrile seizures (0-1 day risk window). Secondary outcomes included febrile seizures (0-7 day risk window) and seizures/convulsions (0-7 day risk window). Seizure outcomes were identified with International Classification of Diseases, Tenth Revision, Clinical Modification (ICD-10-CM) codes in any diagnosis position in the inpatient and emergency department settings. Febrile seizures were identified using ICD-10-CM codes R56.00 and R56.01; seizures/convulsions were identified using ICD-10-CM codes R56.00, R56.01, and R56.9. Incident seizure outcomes were defined as the first recorded seizure event for an individual during the observation period following the exposure, with no previously identified outcomes in the 42-day clean interval prior to outcome.

### Statistical Analysis

We characterized all eligible vaccinees as well the subset of vaccinees with outcomes, including stratification by age, sex, U.S. region, and concomitant vaccination status. We used a conditional Poisson regression model to compare the febrile seizure rates in risk and control intervals and estimate the incidence rate ratio (IRR). From the conditional Poisson regression model, we derived the risk difference (RD) per 100,000 vaccinations and per 100,000 person-years.^9^ We performed meta-analyses to pool results from all three databases, allowing for higher precision and statistical power. Meta-analyses were prespecified as primary and individual database results were specified as secondary. We conducted random-effects meta-analysis to account for between-study heterogeneity across multiple data sources and fixed-effect meta-analysis to address concerns that the random-effects method may not perform well when the number of studies is small. Random-effects meta-analyses are reported in text. We used Cochran’s Q statistic and Higgins & Thompson’s I^2^ statistic to assess between-study heterogeneity.^10^

We conducted a sensitivity analysis restricting to enrollees without concomitant MMR/MMRV (Measles, Mumps, Rubella/Measles, Mumps, Rubella, Varicella), DTaP (Diphtheria, Tetanus, Pertussis), PCV13 (Pneumococcal Conjugate Vaccine), or influenza vaccination on the same day as COVID-19 vaccination. We also adjusted for potential time-varying confounding due to the length of the observation period by adjusting for baseline febrile seizure risk estimated from within-study background rates. We conducted a sensitivity analysis implementing the Farrington adjustment for event-dependent exposure.^11^ Finally, we conducted a sensitivity analysis adjusting for the PPV of our primary outcome using the Schenker and Rubin method and the PPV derived by the Sentinel initiative (PPV 91%; 95% CI 85-95%).^11,12^ Medical record review of the cases identified in the present study is ongoing.

All analyses were conducted using R (version 4.1.2, R Foundation for Statistical Computing) and SAS v. 9.4 (SAS Institute Inc., Cary, NC, United States).

## RESULTS

### Primary Outcome

#### Descriptive Analyses

There were 288,754 BNT162b2 and 192,540 mRNA-1273 vaccinations observed among 163,733 and 110,126 enrollees, respectively. In the primary analysis, 88 cases of febrile seizures were observed following vaccination with BNT162b2, of which 7 cases were observed during the risk interval (days 0-1) and 81 were observed during the control interval (days 8-63) (Table 1). There were 67 cases of febrile seizures observed following vaccination with mRNA-1273, of which 10 cases were observed during the risk interval (days 0-1) and 57 during the control interval (days 8-63).

**Table 1.**
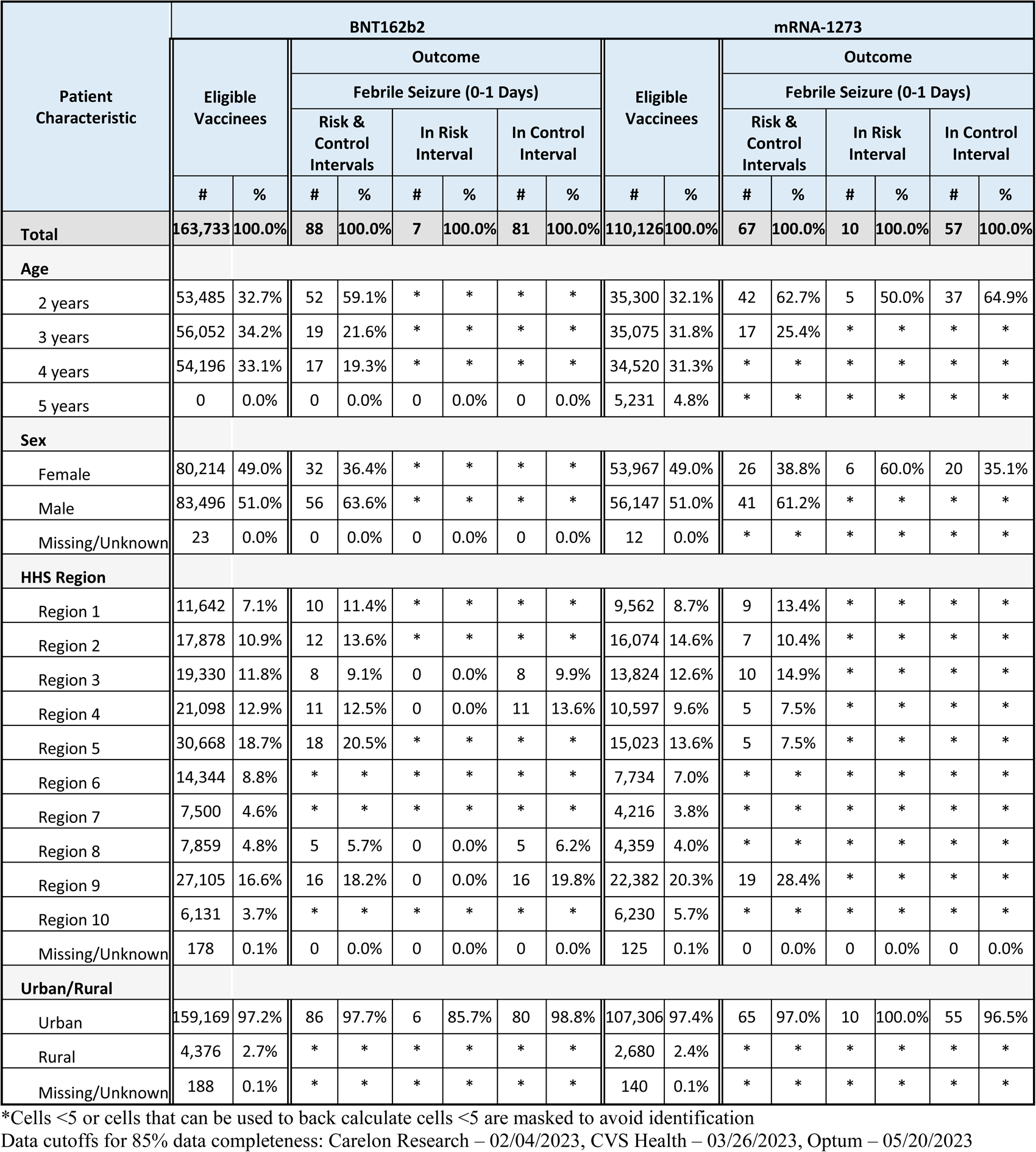
Characteristics of Vaccinated Patients and Patients With Febrile Seizures Aged 2-4 Years (BNT162b2) and 2-5 Years (mRNA-1273), Combined Across Databases.

Table 1 provides a summary of the baseline characteristics of the eligible vaccinated population as well as the population who experienced an outcome in the primary analysis. Among all eligible vaccinees, sex was evenly distributed and age was generally evenly distributed. However, the population experiencing outcomes skewed younger than the ‘all eligible vaccinees’ population, with a greater proportion of cases observed in those aged 2 years compared to all other age groups.

#### Inferential Analyses

Meta-analyzed IRRs for primary and sensitivity analyses are presented in Figure 1 and Table 2. The primary meta-analysis found more than twice the incidence of the primary outcome febrile seizure 0-1 days following the mRNA-1273 vaccine compared to control interval (IRR: 2.52, 95% CI: 1.35 to 4.69) with low between-database heterogeneity (I^2^=0%, 95% uncertainty interval 0-78%; Cochran’s Q test p-value = 0.62). For the BNT162b2 vaccination, the IRR was elevated but not statistically significant (IRR: 1.41, 95%CI: 0.48 to 4.11) with moderate between-database heterogeneity (I^2^=51%, 95% uncertainty interval 0-86%; Cochran’s Q test p-value = 0.13). We estimated a RD of 3.22 (95%CI -0.31 to 6.75) febrile seizure events per 100,000 mRNA-1273 vaccinations, and -0.25 (95%CI -2.75 to 2.24) febrile seizure events per 100,000 BNT162b2 vaccinations (Table 3).

**Figure 1.**
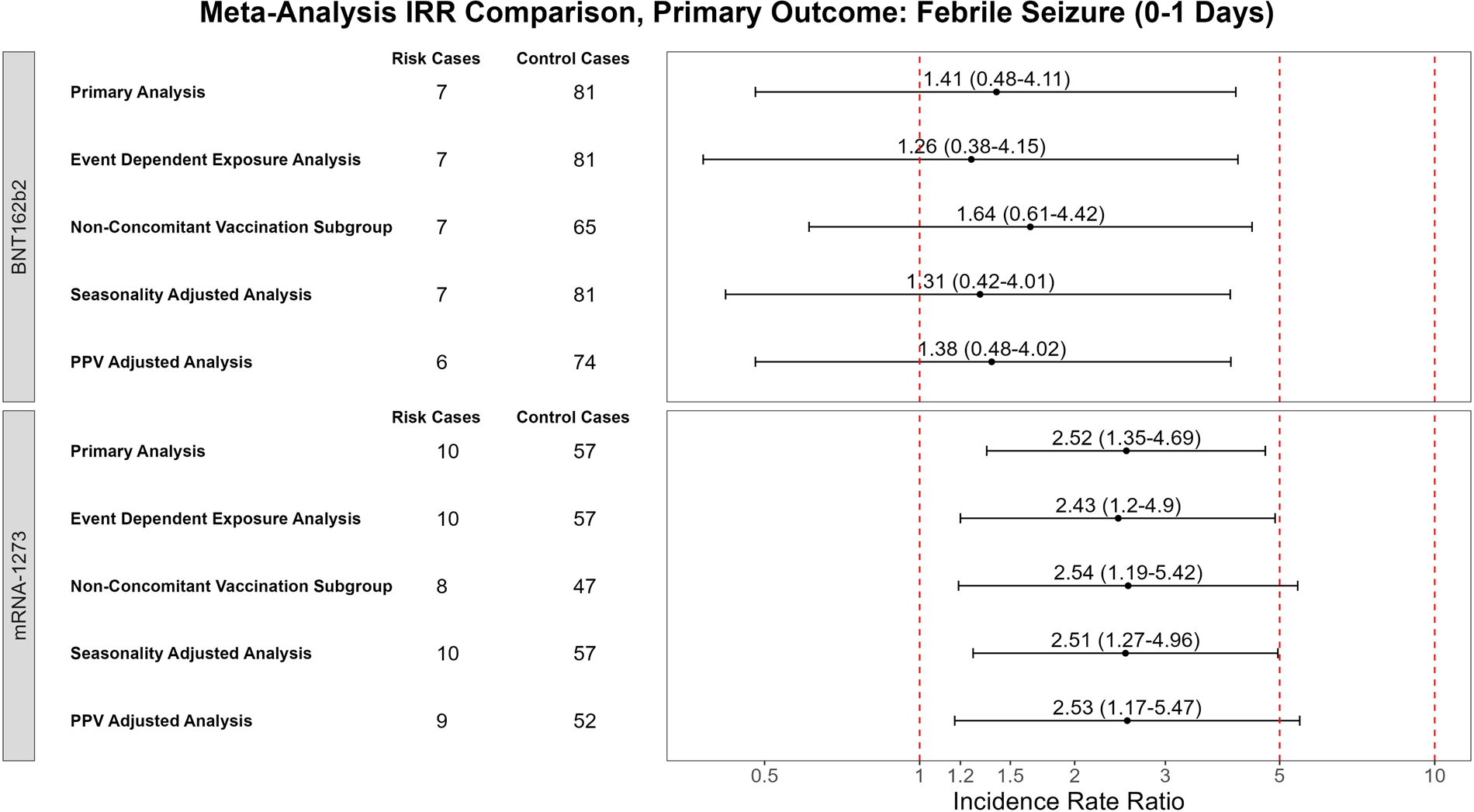
Incidence Rate Ratios for Primary and Sensitivity Random-Effects Meta-Analyses, Febrile Seizures (0-1 Days) Outcome illustrates incidence rate ratios and 95% confidence intervals across analyses for direct comparison.

**Table 2.** Incidence Rate Ratios for Primary and Sensitivity Meta-Analyses, Febrile Seizure (0-1 Days) Outcome.

| Analysis | Brand | Count of Cases | | Count of Days | | Random Effects Meta-Analysis | | | Fixed Effects Meta-Analysis | | | Between-Study Var. ($\tau^2$) | | | Cochran's Q | | I<sup>2</sup> | | |
| --- | --- | --- | --- | --- | --- | --- | --- | --- | --- | --- | --- | --- | --- | --- | --- | --- | --- | --- | --- |
|  |  | Risk | Control | Risk | Control | RR | 95% CI |  | RR | 95% CI |  | Est. | 95% CI |  | Est. | P-Value | Est. | 95% CI |  |
| Primary Analysis | BNT-162b2 | 7 | 81 | 296 | 4,318 | 1.41 | 0.48 | 4.11 | 1.85 | 0.94 | 3.62 | 0.43 | 0.00 | 36.07 | 4.07 | 0.13 | 0.51 | 0.00 | 0.86 |
| Farrington-Adjusted |  | 7 | 81 | 296 | 4,318 | 1.26 | 0.38 | 4.15 | 1.51 | 0.67 | 3.42 | 0.54 | 0.00 | 45.33 | 3.91 | 0.14 | 0.49 | 0.00 | 0.85 |
| Non-Concomitant Vaccines |  | 7 | 65 | 252 | 3,547 | 1.64 | 0.61 | 4.42 | 1.81 | 0.81 | 4.03 | 0.23 | 0.00 | 31.36 | 2.88 | 0.24 | 0.31 | 0.00 | 0.93 |
| Seasonality Adjusted |  | 7 | 81 | 296 | 4,318 | 1.31 | 0.42 | 4.01 | 1.54 | 0.69 | 3.41 | 0.44 | 0.00 | 39.27 | 3.71 | 0.16 | 0.46 | 0.00 | 0.84 |
| PPV-Adjusted |  | 6 | 74 | 269 | 3,929 | 1.38 | 0.48 | 4.02 | 1.52 | 0.63 | 3.65 | 0.26 | 0.00 | 34.13 | 2.87 | 0.24 | 0.30 | 0.00 | 0.93 |
| Primary Analysis | mRNA-1273 | 10 | 57 | 234 | 3,319 | **2.52** | **1.35** | **4.69** | **2.52** | **1.35** | **4.69** | 0.00 | 0.00 | 5.43 | 0.95 | 0.62 | 0.00 | 0.00 | 0.78 |
| Farrington-Adjusted |  | 10 | 57 | 234 | 3,319 | **2.43** | **1.20** | **4.90** | **2.43** | **1.20** | **4.90** | 0.00 | 0.00 | 3.23 | 0.51 | 0.77 | 0.00 | 0.00 | 0.59 |
| Non-Concomitant Vaccines |  | 8 | 47 | 194 | 2,738 | **2.54** | **1.19** | **5.42** | **2.54** | **1.19** | **5.42** | 0.00 | 0.00 | 9.46 | 1.03 | 0.60 | 0.00 | 0.00 | 0.80 |
| Seasonality Adjusted |  | 10 | 57 | 234 | 3,319 | **2.51** | **1.27** | **4.96** | **2.51** | **1.27** | **4.96** | 0.00 | 0.00 | 4.79 | 0.83 | 0.66 | 0.00 | 0.00 | 0.90 |
| PPV-Adjusted |  | 9 | 52 | 213 | 3,017 | **2.53** | **1.17** | **5.47** | **2.53** | **1.17** | **5.47** | 0.00 | 0.00 | 5.86 | 0.80 | 0.67 | 0.00 | 0.00 | 0.90 |

**Table 3.** Risk Difference (RD) Estimates for Primary and Sensitivity Meta-Analyses, Febrile Seizures (0-1 Days Outcome)

| Analysis | Brand | Eligible<br>Vaccinations | Eligible<br>Person-Years | Random-Effects Meta-Analysis |  |  |  |  |  | Fixed Effects Meta-Analysis |  |  |  |  |  |
| --- | --- | --- | --- | --- | --- | --- | --- | --- | --- | --- | --- | --- | --- | --- | --- |
|  |  |  |  | RD Per 100k Doses |  |  | RD Per 100k Person-Years |  |  | RD Per 100k Doses |  |  | RD Per 100k Person-Years |  |  |
|  |  |  |  | AR | 95% CI |  | AR | 95% CI |  | AR | 95% CI |  | AR | 95% CI |  |
| Primary Analysis | BNT-162b2 | 288,754 | 1,580.0 | -0.25 | -2.75 | 2.24 | -46.01 | -501.88 | 409.86 | -0.66 | -2.26 | 0.94 | -120.57 | -413.04 | 171.91 |
| Farrington-Adjusted |  | 288,754 | 1,580.0 | 0.00 | -2.55 | 2.55 | 0.25 | -465.60 | 466.10 | -0.77 | -1.83 | 0.28 | -141.55 | -333.91 | 50.81 |
| Non-Concomitant Vaccines |  | 232,484 | 1,272.4 | 0.29 | -2.18 | 2.77 | 53.03 | -399.18 | 505.24 | -0.21 | -1.81 | 1.38 | -38.93 | -330.27 | 252.40 |
| Seasonality Adjusted |  | 289,115 | 1,582.0 | -0.09 | -2.38 | 2.19 | -17.10 | -434.33 | 400.12 | -0.66 | -1.95 | 0.63 | -120.69 | -356.99 | 115.61 |
| PPV-Adjusted |  | 289,115 | 1,582.0 | -0.18 | -2.02 | 1.66 | -32.25 | -368.72 | 304.22 | -0.49 | -1.77 | 0.79 | -89.93 | -323.52 | 143.65 |
| Primary Analysis | mRNA-1273 | 192,540 | 1,053.6 | 3.22 | -0.31 | 6.75 | 587.81 | -57.35 | 1232.96 | 3.22 | -0.31 | 6.75 | 587.81 | -57.35 | 1232.96 |
| Farrington-Adjusted |  | 192,540 | 1,053.6 | **2.49** | **0.19** | **4.80** | **455.55** | **33.94** | **877.15** | **2.49** | **0.19** | **4.80** | **455.55** | **33.94** | **877.15** |
| Non-Concomitant Vaccines |  | 159,157 | 871.1 | 2.35 | -0.66 | 5.36 | 429.54 | -119.83 | 978.90 | 2.35 | -0.66 | 5.36 | 429.54 | -119.83 | 978.90 |
| Seasonality Adjusted |  | 193,188 | 1,057.1 | 2.58 | -0.21 | 5.38 | 472.23 | -37.94 | 982.40 | 2.58 | -0.21 | 5.38 | 472.23 | -37.94 | 982.40 |
| PPV-Adjusted |  | 193,188 | 1,057.1 | 2.33 | -0.50 | 5.17 | 426.12 | -92.18 | 944.43 | 2.33 | -0.50 | 5.17 | 426.12 | -92.18 | 944.43 |

Sensitivity analysis results were consistent in direction and magnitude with the primary analysis (Figure 1). Farrington-adjusted analyses yielded an IRR of 2.43 (95% CI: 1.35 to 4.69) following mRNA-1273 and 1.26 (95%CI: 0.38 to 4.15) following BNT162b2. Analysis on the subset of the population without concomitant vaccinations yielded an IRR of 2.54 (95% CI: 1.19 to 5.42) following mRNA-1273 and 1.64 (95%CI: 0.61 to 4.42) following BNT162b2. Analyses adjusting for seasonality of seizures resulted in an IRR of 2.51 (95% CI: 1.27 to 4.96) following mRNA-1273 and 1.31 (95%CI: 0.42 to 4.01) following BNT162b2. Finally, PPV-adjusted analyses showed an IRR 2.53 (95% CI: 1.17 to 5.47) following mRNA-1273 and 1.38 (95%CI: 0.48 to 4.02) following BNT162b2.

IRRs from analyses in individual databases are displayed in eFigure 1. IRRs following mRNA1273 were consistently elevated across databases, though only the results in the CVS Health database were significant (Optum IRR=2.19, 95% CI: 0.49 to 9.85; Carelon IRR=1.68, 95% CI: 0.5 to 5.61; CVS IRR=3.59, 95% CI: 1.33 to 9.67). IRRs showed more variation between databases following BNT16b2 vaccination, with significant elevated incidence rate ratios observed only in the Optum database (Optum IRR=3.07, 95% CI: 1.04 to 9.10; Carelon IRR=0.43, 95% CI: 0.06 to 3.17; CVS IRR=0.96, 95% CI: 0.23 to 4.03). Sensitivity analysis results from individual databases were largely consistent with the primary analysis results from the databases.

#### Secondary Outcomes

In the secondary outcomes using a longer risk interval (0-7 days), there were 103 cases of febrile seizures and 135 cases of seizures/convulsions observed following vaccination with BNT162b2, of which 22 cases of febrile seizures and 32 cases of seizures/convulsions were observed during the risk interval. There were 78 cases of febrile seizures and 106 cases of seizures/convulsions observed following vaccination with mRNA-1273, of which 21 cases of febrile seizures and 28 cases of seizures/convulsions were observed during the risk interval.

No statistically significant results were observed in the meta-analysis nor in individual databases for the secondary outcomes (eFigures 2-3). Additionally, no statistically significant results were observed for the secondary outcome populations in any of the analyses with adjustments.

## DISCUSSION

In our study of large cohorts aged 2-5 years in three U.S. commercial health insurance databases, our primary meta-analysis identified a statistically significant increased incidence of febrile seizures among ages 2-4 years in the 0-1 days following mRNA-1273 vaccination. No significantly increased incidence was identified following BNT162b2 vaccination among ages 2-5 years. Database specific results showed increased incidence of febrile seizure immediately following BNT162b2 vaccination in the Optum database and for the mRNA-1273 vaccine in the CVS Health database. No significant increased incidence was identified for secondary outcomes including febrile seizures in the 0-7 days following vaccination and seizures/convulsions in the 0-7 day risk window.

Though our primary meta-analysis showed a relative risk near 2.5 for febrile seizures immediately following mRNA-1273 vaccination, we estimated the risk difference of febrile seizures following vaccination to be small. In the 0-1 day risk interval following over 190,000 eligible mRNA-1273 vaccinations only 10 febrile seizure cases were observed. Our analysis found a risk difference of approximately 3 additional cases of febrile seizures per 100,000 mRNA-1273 vaccinations, though the range from the confidence interval for the estimate included the potential for fewer than zero febrile seizure events per 100,000 vaccinations. The uncertainty in our estimates reflected the rarity of the event. We did not find a statistically significant estimate following BNT162b2 vaccination. Furthermore, there was a lack of consistency in directionality of the incidence rate ratio and attributable risk estimates following BNT1262. This uncertainty is likely due to variation from small case counts and a small number of databases contributing to the meta-analysis.

Our meta-analysis results showed a significantly increased incidence following mRNA-1273 but not following BNT162b2 vaccination. The difference in formulation of the two vaccinations may yield a different immune reaction. The mRNA-1273 vaccination contains 25 µg of antigen per 0.25mL dose, while the BNT162b2 vaccination contains 3 µg of antigen per 0.3mL dose.

Other studies from clinical trials, active surveillance systems, and passive surveillance systems found febrile seizures to be rare and did not report any increased risk for febrile seizures following COVID-19 vaccination in children under 5 years of age. Initial vaccine safety data indicate that among young children, seizures/convulsions following COVID-19 mRNA vaccines are rare: A clinical trial of 3,013 vaccine recipients in children aged 6 months to 4 years reported 5 febrile convulsions cases and only one of those (in a 6-month-old participant) was considered possibly related to the BNT162b2 vaccination or may also have been caused by a concurrent viral infection.^13^ In an analysis of the passive surveillance Vaccine Adverse Event Reporting System (VAERS) data, only 8 seizures were identified following approximately one million COVID-19 mRNA vaccinations through August 2022 in the age group 6 months to 5 years. Six of the 8 seizures were afebrile on medical evaluation.^14^ In CBER’s NRS of the bivalent formulations of the mRNA vaccines authorized in December 2022 for the youngest age groups, no seizures/convulsions outcomes were identified in vaccinees aged 2-5 among 611 person-years of follow up.

It is important to note that febrile seizures occur at rate of up to 5% in young children.^15^ The risk difference we found of 3 additional cases of febrile seizures per 100,000 mRNA-1273 vaccinations is not large compared to some other vaccines and combinations of vaccines which may carry a small risk of at most 30 febrile seizures in 100,000 children vaccinated.

Our study has several strengths. First, our study included a large, geographically diverse population from three US commercial health insurance databases. Our use of a self-controlled study design inherently adjusted for time-invariant confounders such as health conditions, socioeconomic status, and demographic characteristics that may introduce bias in other study designs using between-individual comparisons. We adjusted for time-varying confounding such as seasonality in our sensitivity analyses. Our study sampled vaccinated cases from health plan and public health authority data and not from individual-level reports of vaccination status and therefore was not subject to bias due to underreporting of vaccination status. Our study used IIS data to supplement claims-based vaccination information thus increasing sample size of SCCS study for these rare outcomes. Finally, our outcome definition was shown to have a high PPV;^16^ therefore, our study was able to minimize potential outcome misclassification bias.

Our study also has some limitations. First, the rarity of the event led to substantial uncertainty in estimates, and our study was not powered to identify very small increases in risks. The small number of outcomes further prevented us from performing dose-specific analyses or further subgroup analyses. We may have omitted some beneficiaries with outcomes due to delayed reporting; however, we selected study end dates to ensure 85% data completeness. Furthermore, we conducted PPV adjustment using the Sentinel Initiative’s PPV estimates as medical record review for this study is still underway for our population. Sentinel Initiative’s PPV estimates may not be generalizable to our population as they were based on ICD-9 codes. Because of the low number of databases in the meta-analysis, the statistics for measuring heterogeneity between studies could be biased. Due to low observed counts and different formulation, this study focused on monovalent mRNA vaccines and did not include bivalent formulations. Finally, though we did adjust for seasonality, there may be additional bias from unmeasured time-varying covariates such as circulating respiratory viruses such as SARS-CoV-2 and influenza.

## Conclusions

Our study found an approximately 2.5 times increased incidence of febrile seizures among those aged 2-4 years in the 0-1 days following monovalent mRNA-1273 vaccination. However, absolute risk was low. Based on the current body of scientific evidence, the safety profile of the monovalent mRNA vaccines remains favorable for use in young children. Safety monitoring continues as new COVID-19 vaccines become available for children. This study was conducted under the FDA Biologics Effectiveness and Safety (BEST) Initiative, which plays a major role in the larger US federal government vaccine safety monitoring efforts and further supports regulatory decision-making regarding COVID-19 vaccines.

## Data Availability

The study protocol was posted previously. Data analyses and related documents can be made available where needed, by contacting the corresponding author. De-identified participant data will not be shared without approval from the data partners

## Abbreviations

BEST: Biologics Effectiveness and Safety
CBER: Center for Biologics Evaluation and Research
CI: Confidence Interval
CPT: Current Procedural Terminology
CVX: IIS code indicating vaccination
DTaP: Diphtheria, Tetanus, Pertussis
EUA: Emergency Use Authorization
FDA: Food and Drug Administration
HCPCS: Healthcare Common Procedure Coding System
ICD-10-CM: International Classification of Diseases, Tenth Revision, Clinical Modification
IIS: Immunization Information System
IRR: Incidence Rate Ratio
MMR/MMRV: Measles, Mumps, Rubella/Measles, Mumps, Rubella, Varicella
mRNA: Messenger ribonucleic acid
NDC: National Drug Codes
NRS: Near Real-Time Surveillance
PCV13: Pneumococcal Conjugate Vaccine
PPV: Positive Predictive Value
RD: Risk Difference
SAE: Systemic Adverse Event
SCCS: Self-Controlled Case Series
U.S.: United States
VAERS: Vaccine Adverse Event Reporting System

## Contributors Statement

Steven A Anderson and Richard A Forshee conceptualized and designed the study, interpreted data and analysis, drafted the manuscript, critically reviewed and revised the manuscript, and provided supervision of the analysis.

Elizabeth R Smith conceptualized and designed the study, interpreted data and analysis, drafted the manuscript, and critically reviewed and revised the manuscript.

Zhiruo Wan, Kamran Kazemi, and Mao Hu conceptualized and designed the study, designed data collection instruments, carried out the data analysis, and critically reviewed and revised the manuscript.

Yoganand Chillarige conceptualized and designed the study, critically reviewed and revised the manuscript, and provided supervision of the analysis.

Kandace L Amend, Alex Secora, Djeneba Audrey Djibo, Jennifer Song, Lauren E Bartlett, John D Seeger, Nandini Selvam, and Cheryl N McMahill-Walraven carried out data analysis and critically reviewed and revised the manuscript.

All authors approved the final manuscript as submitted and agree to be accountable for all aspects of the work.

## Acknowledgements

Bowen Chen, Vincent Varvaro, Xi Li of Acumen, LLC; Shiva Vojjala, Ramya Avula, Shiva Chaudhary, Shanthi P Sagare, Ramin Riahi, Brian Greenwald, Mia Si, Dianna Hayden, and Grace Stockbower of Carelon Research; Michael Goodman, Ken Revett, Ruth Weed, and Nerissa Williams of IQVIA; Lauren Peetluk, PhD, MPH, Elizabeth J. Bell, PhD, MPH, Wafa Tarazi, PhD, MHPA, Alexandra Stone, PhD, MS of Optum; and Anne Marie Kline MS, CHES, Nancy B. Shaik, BS, Ana M Martinez-Baquero, MA, Vaibhav Sharma, MS, Smita Bhatia, MCA, Yi Liu, MDA, MS, Xun Zhang, MMS, Eugenio Abente, PhD, Jonathan P Deshazo, PhD, MPH, Aparna Srikanti, MSc, Ralph Webber, BS, Charlalynn Harris, PhD, MPH, Wuan M Head, BA, Harpreet Kaur Dhillon, MCA, Carla Brannan, BA, Jack Dimpel, and James Dunlap of CVS Health for their assistance with data validation, analysis, and project coordination.

## Figures Legend

Figure e1. Incidence Rate Ratios, Random Effects Meta-Analysis and Individual Database Results, Primary Outcome: Febrile Seizure (0-1 Day Risk Interval) illustrates incidence rate ratios and 95% confidence intervals across individual databases for all analyses of the primary outcome.

eFigure 2. Incidence Rate Ratios, Random Effects Meta-Analysis and Individual Database Results, Secondary Outcome: Febrile Seizure (0-7 Day Risk Interval) illustrates incidence rate ratios and 95% confidence intervals across individual databases for all analyses of the secondary febrile seizure outcome.

eFigure 3. Incidence Rate Ratios, Random Effects Meta-Analysis and Individual Database Results, Secondary Outcome: Seizure (0-7 Day Risk Interval) illustrates incidence rate ratios and 95% confidence intervals across individual databases for all analyses of the secondary seizure/convulsion outcome.

