## Supplement for "Febrile Seizure Risk Following Monovalent COVID-19 mRNA Vaccination in US Children Aged 2-5 Years"

### eTable 1. Database Characteristics

| **Database** | **Description** | **Claims Type** | **Update frequency** | **Data Lag, Time to 80% Completeness** | **Average No. Enrollees aged 6 months-4 years (2020-2023) ^#^** | **No. IIS Jurisdictions Incorporated into Analysis** | **Study End Date** |
| --- | --- | --- | --- | --- | --- | --- | --- |
| CVS Health | CVS Health transforms Aetna health plan enrollment, demographic, and medical and drug claims data, for individuals enrolled from January 2018 forward. The Aetna data includes commercial, including Affordable Care act (ACA) Marketplace, and Medicare Advantage health plans into a patient-centered, comprehensive Common Data Model (CDM). | Fully Adjudicated | Monthly | ~ 3-4 months for IP claims, 2-3 months for OP claims, and 1-2 months for professional claims | 6 months-4 years: > 1.08m | 22 | 3/26/2023 |
| Optum Pre-adjudicated Claims | The Optum data includes enrollment, prescription drug and pre-adjudicated hospital and physician health insurance claims. The pre-adjudicated claims database includes claims for privately insured. Hospital and physician claims undergo initial processing on a daily basis from a large number of providers across the US who accept patients with health insurance. | Pre-Adjudicated | Once every two weeks | ~ 1-2 months for IP, OP, and professional claims | 6 months-4 years: > 839k | 20 | 5/20/2023 |
| Carelon Research | Carelon Research is a wholly-owned, research subsidiary of Elevance Health, Inc., a holding company owning several large US health plans associated with Anthem Blue Cross Blue Shield. Carelon Research has a US population database including individually insured by commercial and Medicare Advantage plans, the Healthcare Integrated Research Database (HIRD), with longitudinal data on health plan enrollees. | Fully Adjudicated | Monthly | ~ 2-3 months for IP claims and 1-2 months for OP and professional claims | 6 months-4 years: > 1.32m | 8 | 2/4/2023 |

### eTable 2. Administrative Codes for COVID-19 Vaccine Administration Used in Claims and IIS databases

| **HCPCS/CPT* Codes** | **CVX** Codes (IIS-Specific)** | **Manufacturer** | **Name** | **Age Group (years)** | **Vaccine Administration Code** | **NDC*** 11 Labeler Product ID (Vial)** | **Dosing Interval** |
| --- | --- | --- | --- | --- | --- | --- | --- |
| 91308 | 219 | Pfizer | Pfizer-BioNTech COVID-19 Vaccine | 6 m- 4 | 0081A (1st dose) | 59267-0078-01 59267-0078-04 | - 21+ days between dose 1 and dose 2 - 28+ days between dose 2 and dose 3 |
|  |  |  |  |  | 0082A (second dose) |  |  |
|  |  |  |  |  | 0083A (third dose) |  |  |
|  |  |  |  |  | 0052A (2^nd^ dose) |  |  |
|  |  |  |  |  | 0053A (3^rd^ dose) |  |  |
|  |  |  |  |  | 0054A (booster dose) |  |  |
|  |  |  |  |  | 0002A (2^nd^ dose) |  |  |
|  |  |  |  |  | 0003A (3^rd^ dose) |  |  |
|  |  |  |  |  | 0004A (booster dose) |  |  |
|  |  |  |  |  | 0002A (2^nd^ dose) |  |  |
|  |  |  |  |  | 0003A (3^rd^ dose) |  |  |
|  |  |  |  |  | 0004A (booster dose) |  |  |
| 91301, 91306, 91309 | 207 | Moderna | Moderna COVID-19 Vaccine | 6 m-17 | 0011A, 0091A (1^st^ dose) | 61434-0043-00  61434-0043-01  80777-0100-11  80777-0100-98  80777-0100-99  80777-0277-05  80777-0279-05  80777-0273-98  80777-0273-10  80777-0273-99  80777-0273-15  80777-0273-98  80777-0273-05  80777-0273-99 | - 28+ days between dose 1 and dose 2 - For immunocompromised, 28 days between dose 1 and dose 2, and 28+ days between dose 2 and dose 3. |
|  |  |  |  |  | 0012A, 0092A  (2^nd^ dose) |  |  |
|  |  |  |  |  | 0013A, 0093A  (3^rd^ dose) |  |  |
|  |  |  |  |  | 0064A, 0094A (booster dose) |  |  |
|  |  |  |  |  | 0042A  (2^nd^ dose) |  |  |

### Figure e1. Incidence Rate Ratios, Random Effects Meta-Analysis and Individual Database Results, Primary Outcome: Febrile Seizure (0-1 Day Risk Interval)

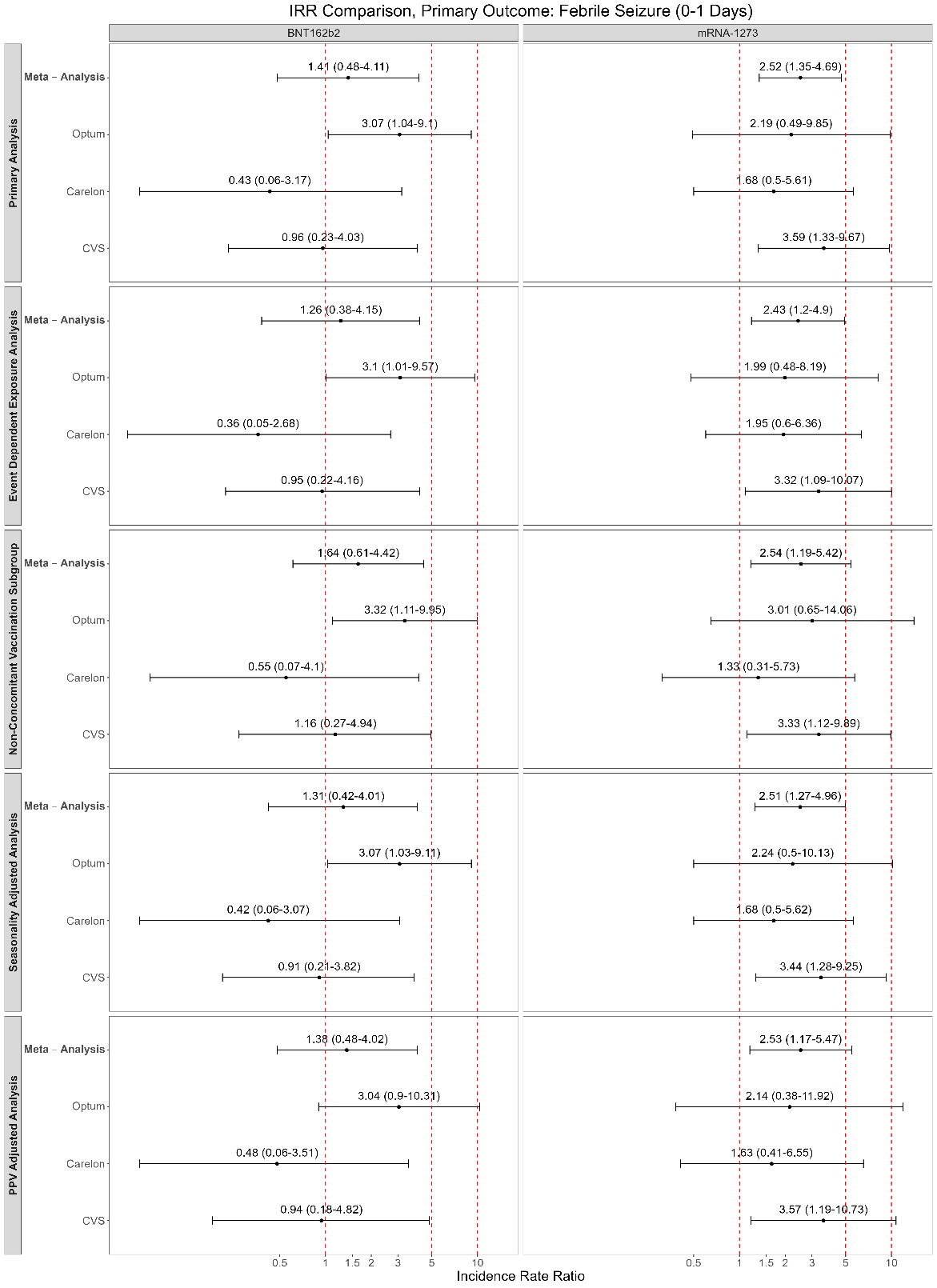

### eFigure 2. Incidence Rate Ratios, Random Effects Meta-Analysis and Individual Database Results, Secondary Outcome: Febrile Seizure (0-7 Day Risk Interval)

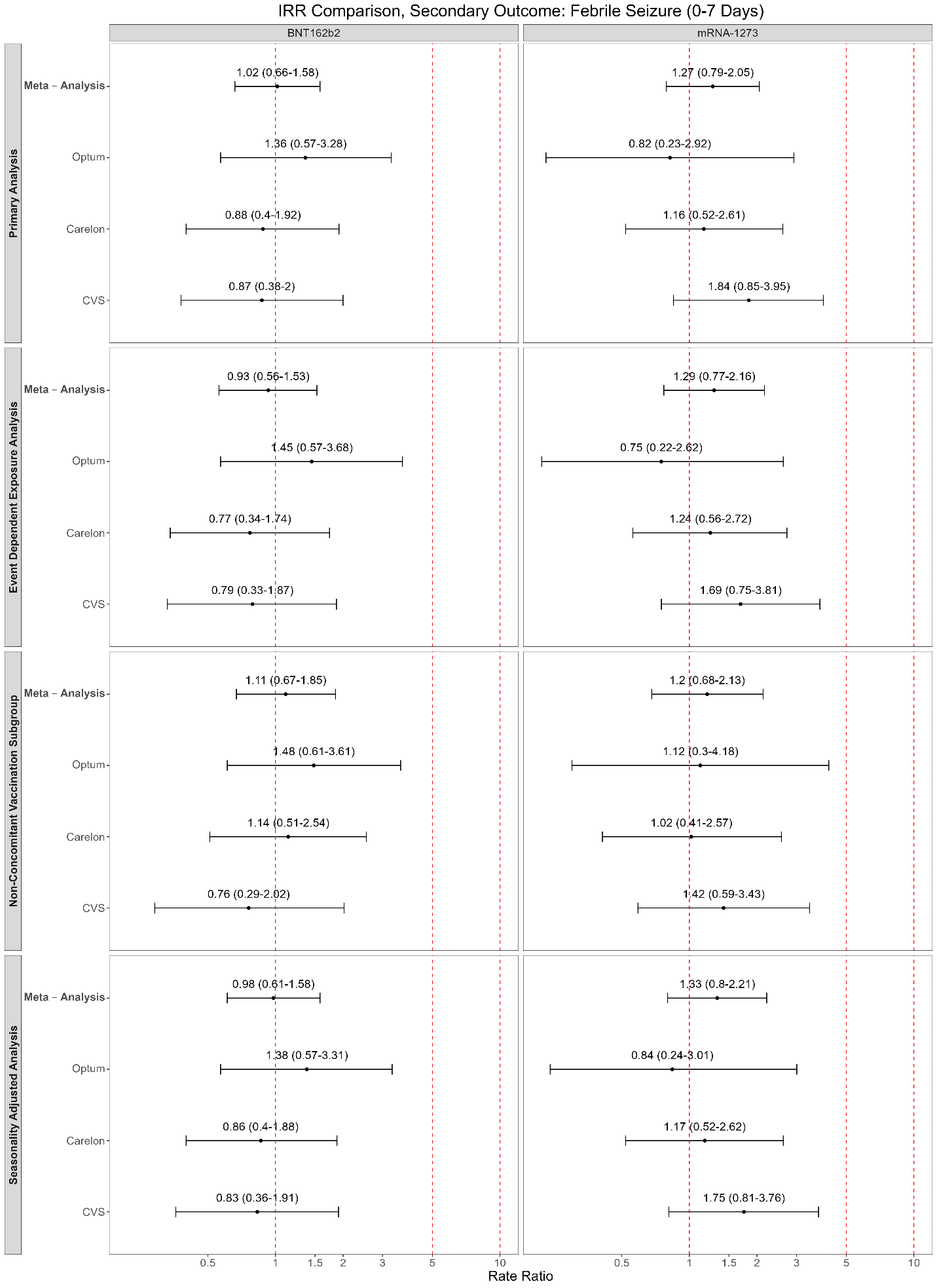

### eFigure 3. Incidence Rate Ratios, Random Effects Meta-Analysis and Individual Database Results, Secondary Outcome: Seizure (0-7 Day Risk Interval)

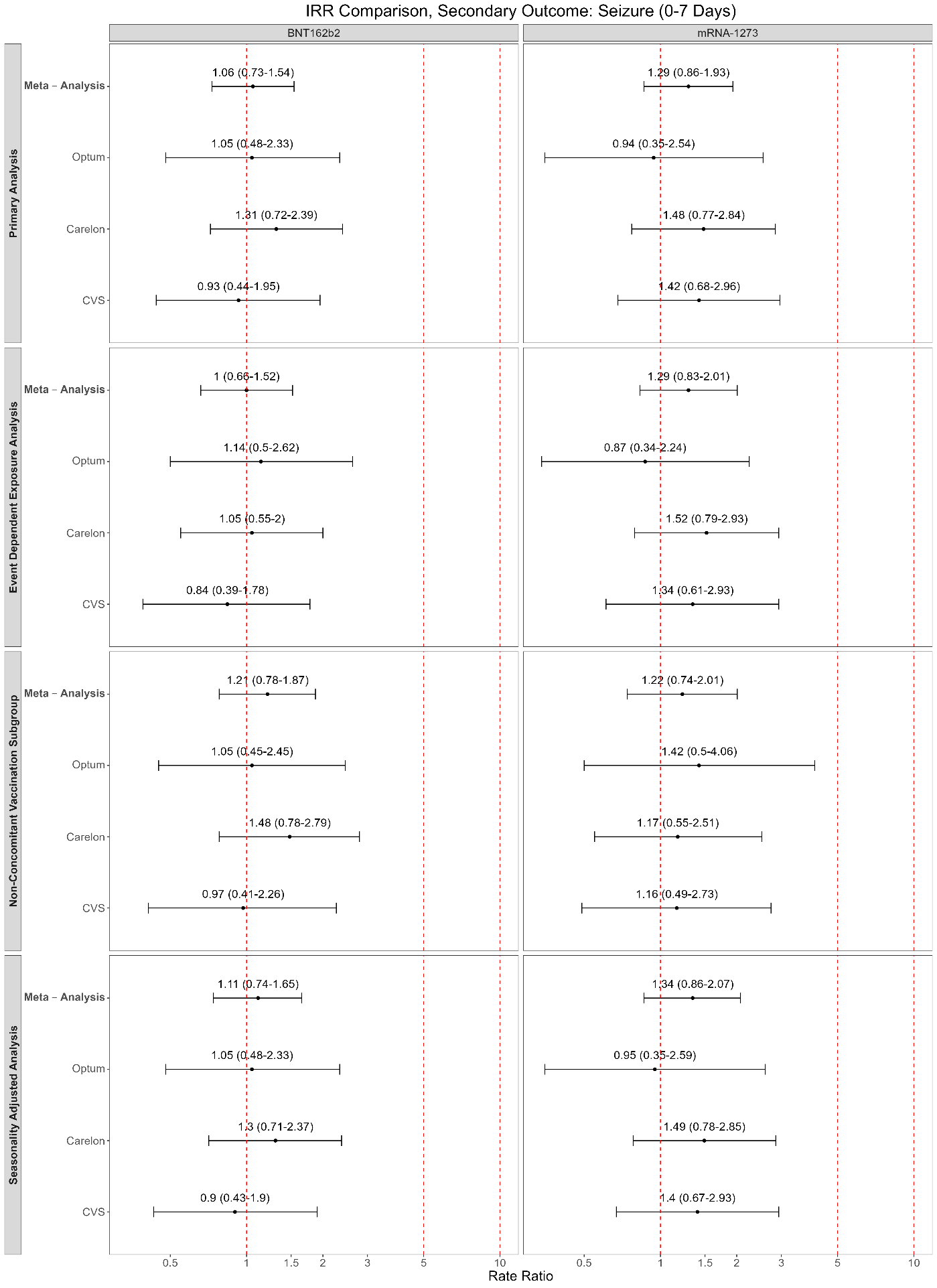
